# Modelling population-wide screening of SARS-CoV-2 infection for containing COVID-19 pandemic in Okinawa, Japan

**DOI:** 10.1101/2020.12.19.20248573

**Authors:** Kazuki Shimizu, Toshikazu Kuniya, Yasuharu Tokuda

**Affiliations:** Department of Health Policy, London School of Economics and Political Science, London, UK; Faculty of Public Health and Policy, London School of Hygiene and Tropical Medicine, London, UK; Graduate School of System Informatics, Kobe University, Kobe, Japan; Muribushi Okinawa Center for Teaching Hospitals, Okinawa, Japan

## Abstract

**Background:** To break the chains of SARS-CoV-2 transmission and contain the coronavirus disease 2019 (COVID-19) pandemic, population-wide testing is practiced in various countries. However, scant research has addressed this topic in Japan.

**Materials and Methods:** In this modelling exercise, we extracted the number of daily reported cases of COVID-19 in Okinawa from October 1 to November 30, 2020 and explored possible scenarios for decreasing COVID-19 incidence by combining population-wide screening and/or social distancing policy.

**Results:** We reveal that permanent lockdown can be replaced by mass testing that mobilizes sufficient target population at an adequate frequency. In addition, solely imposing a circuit breaker will not bring a favorable outcome in the long-term, and mass testing presents implications for minimizing a period of lockdown.

**Discussion:** Our results highlight the importance of incentivizing citizens to join the frequent testing and ensure their appropriate isolation. To contain the COVID-19 pandemic, rigorous investment in public health is manifestly vital.

## Introduction

To combat against the surge of coronavirus disease 2019 (COVID-19) cases and break chains of SARS-CoV-2 transmission, many countries imposed various non-pharmaceutical interventions (NPIs) in 2020. Especially, an unprecedented lockdown of cities, regions, and countries highly contributed to suppress the virus [1]. Although the economic loss and social burden on citizens, especially vulnerable populations, were critically featured, and there has been a dispute on how to leverage socio-economic activities with infection control measures in many countries that employed suppression or mitigation strategies, it becomes evident that economic activities were subdued by social distancing policy, even at a voluntary basis [2]. On the contrary, some countries or regions in the Western Pacific Region, such as China, Taiwan, Vietnam or New Zealand, advanced for aggressively suppressing or eliminating COVID-19, and have started to boost their socio-economic activities [3]. Australia is currently at the final phase for eliminating community-acquired infection under its aggressive suppression strategy.

To contain the COVID-19 pandemic, symptom-based strategy is not feasible due to its large proportion of asymptomatic infections [4], and many countries rapidly ramped up the testing capacity, expanded testing targets, and arranged logistics for rigorous contact-tracing and isolation. There has been an argument that weekly testing for the entire population, followed by contact-tracing and quarantine, can break chains of transmission within a few weeks [5], and population-wide testing has been practiced in various ways [6]. China utilized a mass testing approach for early containment [7,8], and some European countries have started to employ it through a novel approach. Slovakia spearheaded for screening over 3.6 million persons, and mass testing with other NPIs successfully reduced the prevalence of SARS-CoV-2 [9]. The United Kingdom firstly kicked off the trial of mass testing in Liverpool by a lateral flow test, and positive cases were followed by confirmatory reverse transcription polymerase chain reaction (RT-PCR) testing [10]. Other countries, such as South Tyrol/Alto Adige in Italy, Austria and Slovenia, has already conducted or will implement similar approaches.

While many Asian-Pacific countries have rigorously contained the COVID-19, Japan has been employing the suppression strategy. While maintaining the reproduction number at least below 1 will be vital in this strategy, the government kept neglecting its evolution [11]. Rather than strengthening additional countermeasures for high-risk environments, imposing social distancing policies and ensuring citizens’ access to testing, the government boosted multiple “go to” campaigns [12]. As a result, the spread of the SARS-CoV-2 virus has been accelerated to the entire nation. Some clusters of COVID-19 have already been reported from remote islands in Hokkaido or Okinawa prefectures.

After lifting the nationwide state of emergency in early May, Okinawa prefecture, whose population amounts to 1.46 million with 148 remote islands, aggressively suppressed the virus, and the number of COVID-19 cases was maintained almost zero in May-June 2020; however, there were insufficient efforts for expanding testing targets to truly eliminate the virus. This, along with the importation from other prefectures and COVID-19 outbreak in U.S. military bases, brought a resurgence in late July, and necessitated local state of emergency in August. After lifting it, the grand strategy has obscurely revised. Now Okinawa has been again facing an increase of COVID-19, and the purpose of this study is to explore possible scenarios for decreasing its incidence by combining population-wide testing and/or social distancing policy.

## Materials and Methods

### Model

In the numerical simulation, we used an SEIQR compartmental model as in [13], in which the susceptible, exposed (asymptomatic infectious), infectious, quarantined and recovered populations are considered. As in [13], we assumed that, for COVID-19, the average incubation period is 5 days [14], the average infectious period is 10 days [15], the average quarantine period is 14 days and the reproduction numbers for asymptomatic and symptomatic infections are 0.44 R and 0.56 R, respectively [16], where R denotes the reproduction number. To estimate R in Okinawa, we applied the least-squares method as in [17] under the assumption that the identification rate is 0.1 to the data of number of daily reported cases of COVID-19 in Okinawa from October 1 to November 30, 2020. We then obtained R=1.22, and the asymptotic and symptomatic infection rates were calculated as in [13]. The sensitivity and specificity in usual diagnosis were assumed as 90% and 100%, respectively [18]. In the pulsed mass testing that screens infectiousness among citizens people, which is described in the next subsection, we assumed a perfect test sensitivity for detecting infectious individuals regardless of their symptoms, specificity and compliance with quarantine, as illustrated in another modelling study of population-wide screening [9].

### Multiple scenarios

We simulated multiple scenarios under the assumption that the pulsed mass testing and lockdown started on December 8, 2020. On the mass testing, we assumed that the testing is periodically carried out per 7/14/30 days to 5/25/50% of the whole population in Okinawa. On the other hand, we also simulated a scenario that the daily self-testing is carried out on weekdays to 1/5/10% of the whole population in Okinawa. Consequently, we obtained four figures (see the next section) for the following scenarios.

i. Mass testing with testing rate 5% per 7/14/30 days;
ii. Mass testing with testing rate 25% per 7/14/30 days;
iii. Mass testing with testing rate 50% per 7/14/30 days;
iv. Daily self-testing with testing rate 1/5/10%.

In addition, in each scenario, we considered three cases of social distancing policies that decreased social contacts by 50%, and the duration of lockdown was classified into three patterns: no lockdown: circuit breaker where the lockdown ends after 4 weeks; and the permanent lockdown.

## Results

Multiple scenarios of mass testing towards 5%. 25%, and 50% of whole population in Okinawa at different intervals are presented in Figures 1-3. The number of daily reported cases will exponentially increase without mass testing or lockdown. While imposing the circuit breaker can curb the epidemic, it is not sufficient to aggressively suppress the virus; therefore, to decrease the number of patients, mass testing at appropriate intervals must be combined. The permanent lockdown will decrease COVID-19 cases, and its period will be shortened by simultaneously implementing mass testing. Figure 1 clearly exhibits that mass testing for 5% of all citizens in Okinawa is not sufficient to curb an increasing number of daily reported cases, even when mass testing is conducted on a weekly basis or combined with circuit breaker (Figure 1).

**Figure 1:**
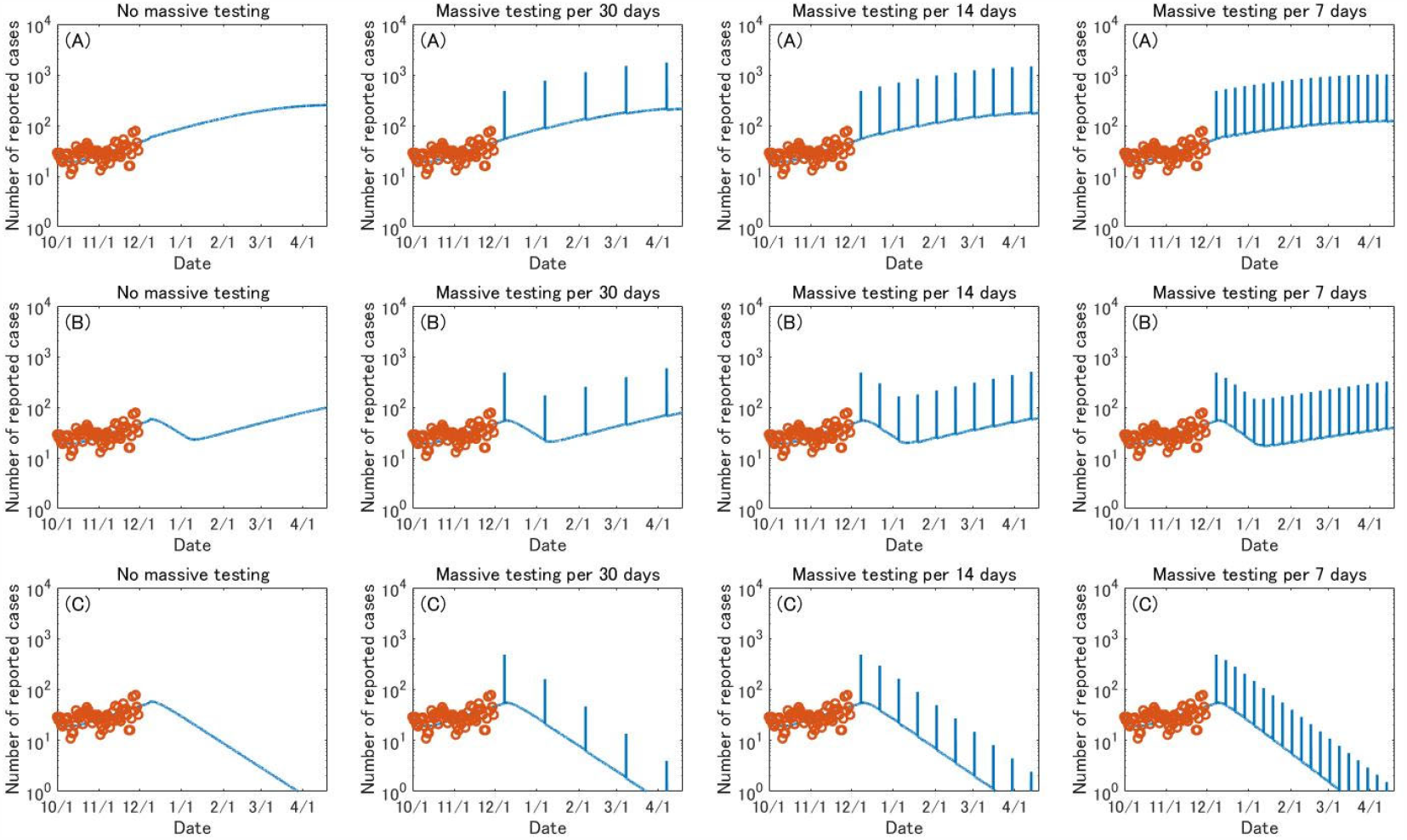
Evolution of daily reported COVID-19 cases when population-wide testing is performed for 5% of all citizens at specific intervals. (A) no lockdown; (B) circuit breaker; (C) permanent lockdown

This tendency slightly changes when 25% of whole population in Okinawa are screened, as presented in Figure 2. When the lockdown is not a policy option, mass testing per 14 days will curb an increasing trend; however, each mass testing continuously reports more than 1,000 cases for a few months. When implementing mass testing per 7 days, an increasing trend will be reversed; however, it must be debatable in terms of health system capacity. Obviously, a frequent testing will hugely contribute to breaking chains of transmission, and vividly decreasing the reported number of COVID-19 per mass testing. Even without lockdown, the number of people tested positive will decrease to below 100 after April. When combined with the circuit breaker, the number will be decreased to below 100 in early February (Figure 2).

**Figure 2:**
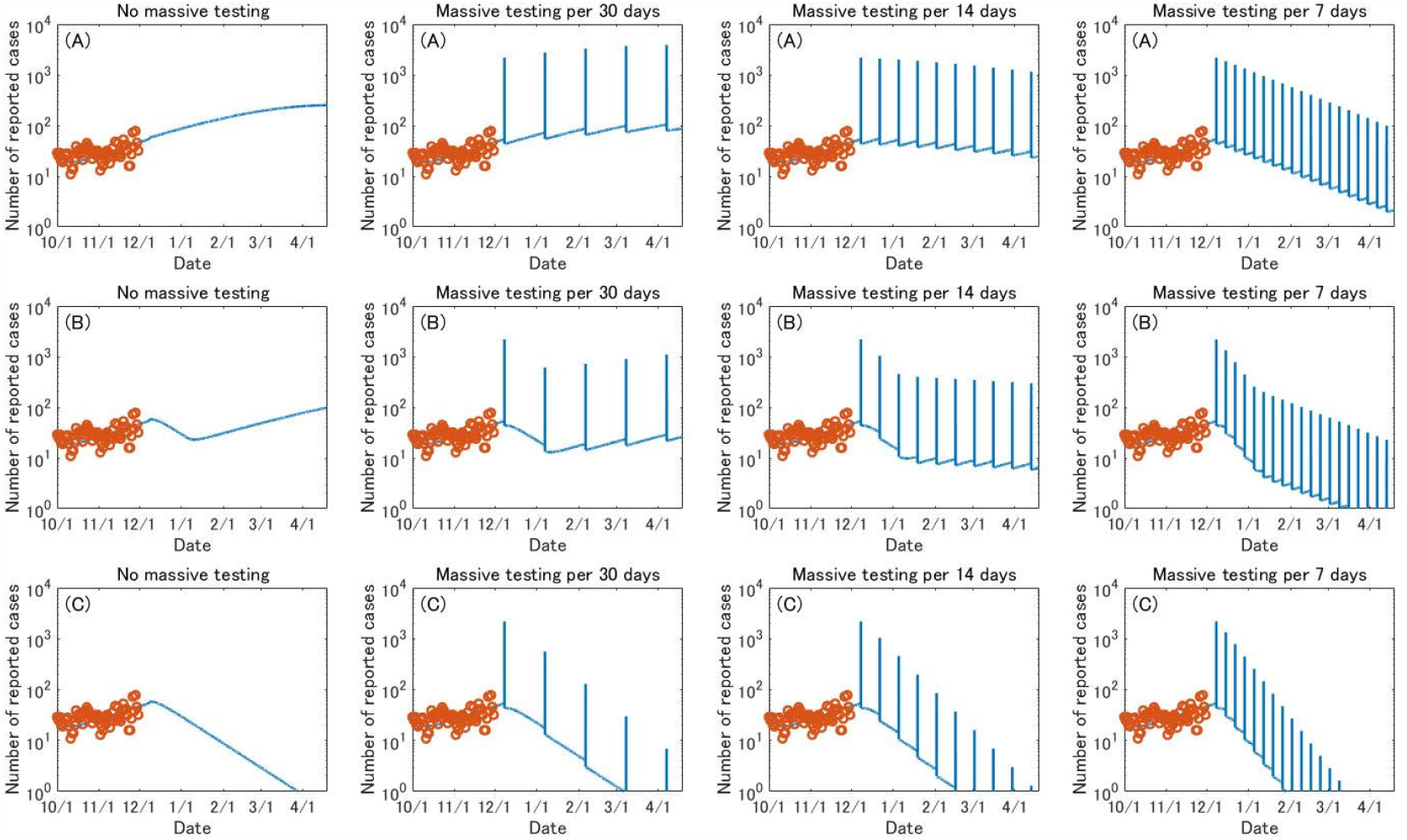
Evolution of daily reported COVID-19 cases when population-wide testing is performed for 25% of all citizens at specific intervals. (A) no lockdown; (B) circuit breaker; (C) permanent lockdown

Figure 3 illustrates scenarios of mass testing that targets 50% of all citizens. Even monthly mass testing will quell an increasing trend of transmission; however, it should be noted that even in April, more than 1,000 cases will be reported. Mass testing per 14 days will truly curb the epidemic, and the decreasing trend will be accelerated by simultaneously implementing mass testing and imposing a circuit breaker. When the weekly mass testing is performed without lockdown, it will promptly impact on crashing transmission dynamics, and record less than 10 cases in late-February. This period can be shortened to early February when combined with circuit breaker. Figure 3A and Figure 3C also suggest that weekly mass testing for 50% of entire population will not only overturn an increasing trend of COVID-19 patients but bring a similar effect that can be gained by simply imposing a permanent lockdown (Figure 3).

**Figure 3:**
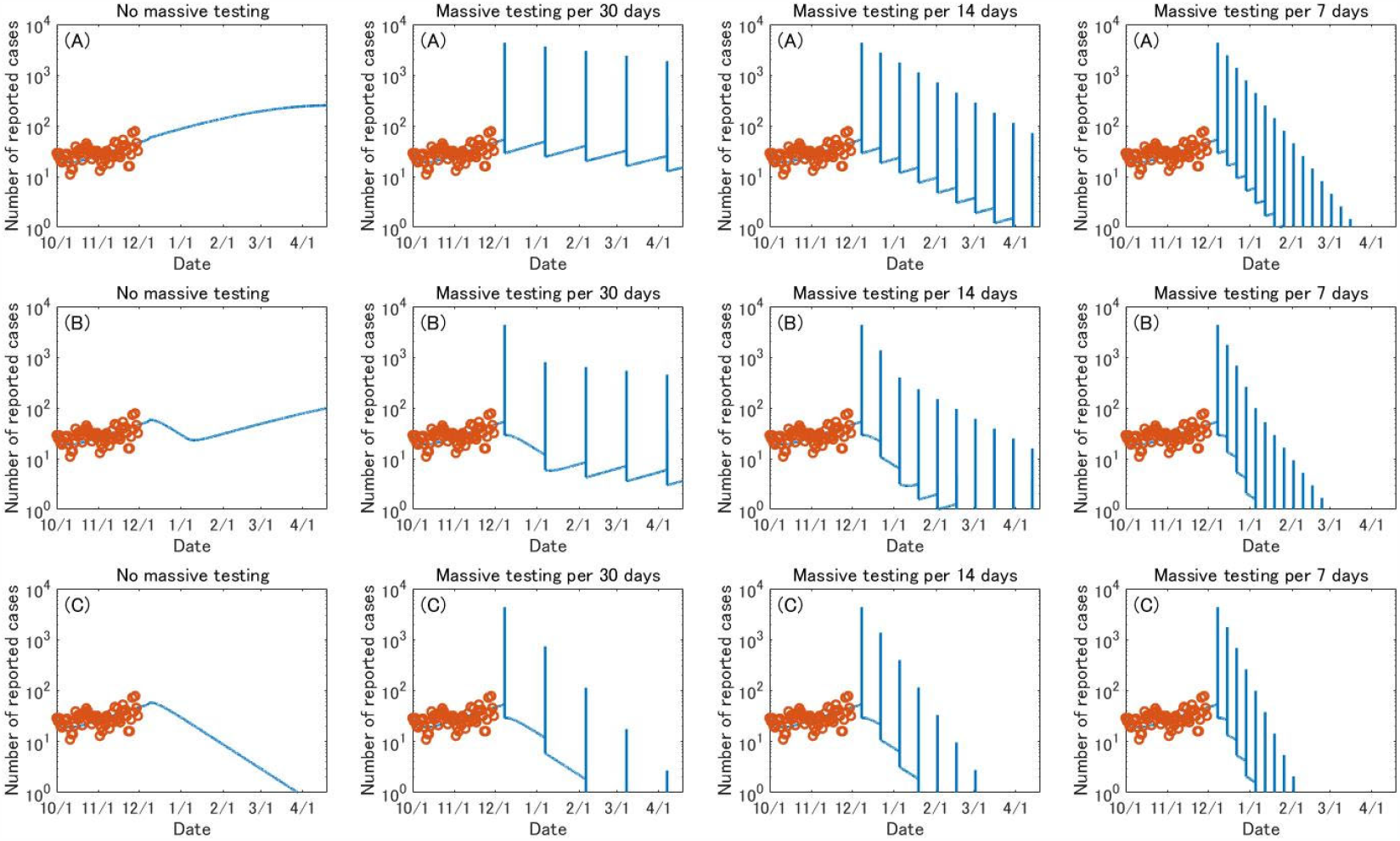
Evolution of daily reported COVID-19 cases when population-wide testing is performed for 50% of all citizens at specific intervals. (A) no lockdown; (B) circuit breaker; (C) permanent lockdown

Figure 4 presents the evolution of daily reported cases when the testing is conducted on a daily basis. Our results show that daily testing towards 1% of all citizens is not sufficient to hammer an increasing trend of daily reported cases without permanent lockdown. When 5% of all citizens are targeted, the number of reported cases will gradually decrease but it will record over 100 cases even in March: therefore, this scenario needs to be carefully considered in terms of health system capacity. When 10% of all citizens were tested on a daily basis, the number of daily reported cases will reach below 10 in late February. When combined with circuit breaker, daily testing towards 1% of all citizens is not enough to turn the tide; but expanding more than 5% of all citizens will be effective to decrease the number of daily reported cases (Figure 4).

**Figure 4:**
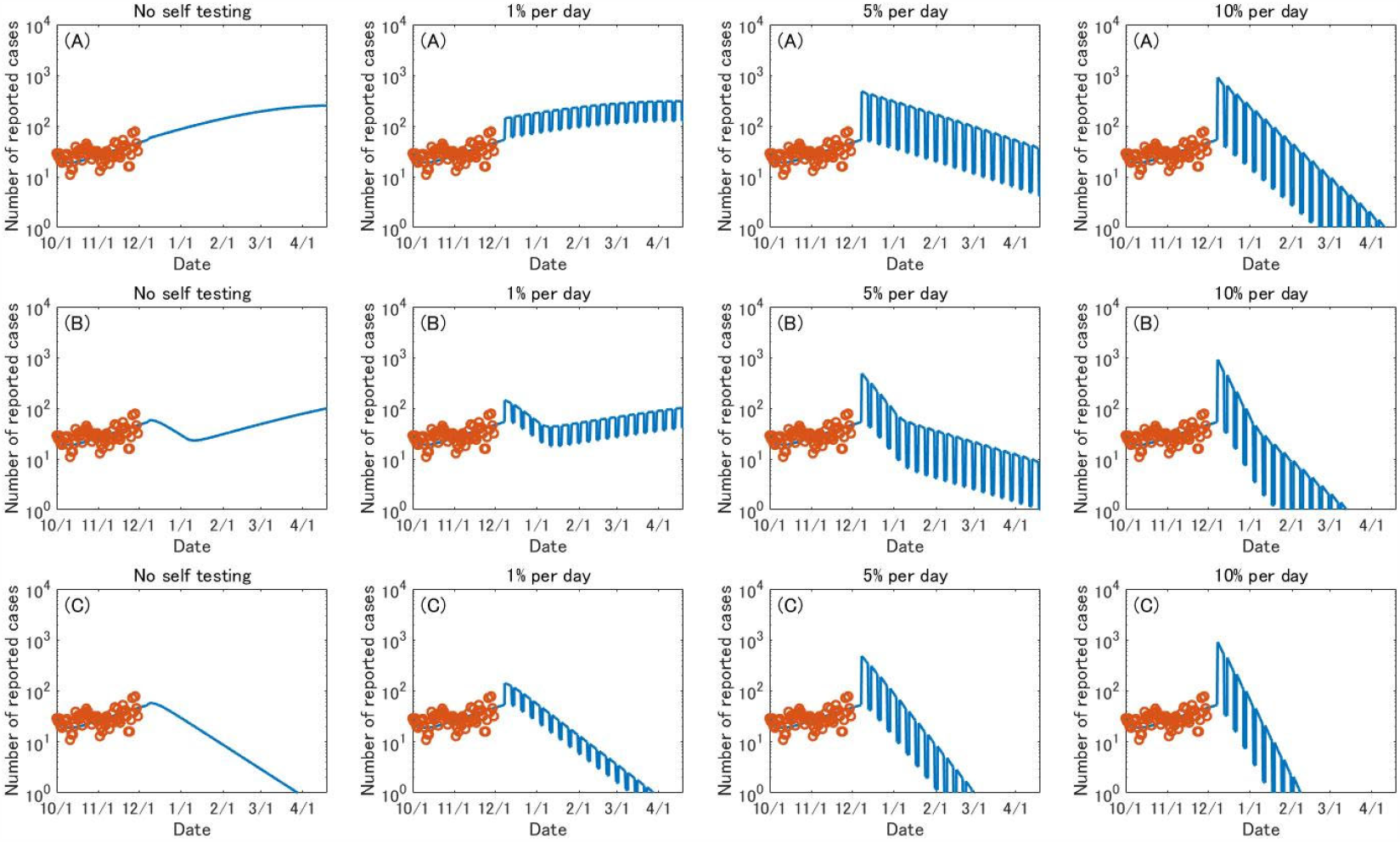
Evolution of daily reported COVID-19 cases when population-wide testing is performed on a daily basis. (A) no lockdown; (B) circuit breaker; (C) permanent lockdown

## Discussion

Our modelling exercise revealed three major findings. First, there is a scenario that permanent lockdown can be averted by population-wide testing. If the mass testing covers 50% of all citizens on a weekly basis, the number of COVID-19 cases will reach nearly zero in mid-March. Even when the project is conducted through daily testing approach and targets 10% of all citizens, it will also contribute to decreasing COVID-19 cases and the new COVID-19 cases will reach nearly zero in early April. As mass testing that targets 5% of all citizens at a weekly basis or daily testing for 1% of all citizens will not change the deteriorating trend, mobilizing sufficient target population will be vital. Second, a frequent mass testing will be crucial to curb the ongoing COVID-19 pandemic in Okinawa, even when the current test-trace-isolate systems are maintained. Monthly screening will not bring a decrease of COVID-19 infections, and an adequate frequency needs to be ensured. Third, mass testing presents implications for minimizing a period of lockdown. To truly reverse an increasing trend of COVID-19, circuit breaker, namely a short-term lockdown, must be combined with population-wide screening for sufficient population. Solely imposing a circuit breaker will not bring a favorable outcome in the long-term.

Our results are in line with other latest evidence. As pre-symptomatic and asymptomatic infections contribute to the whole transmission of SARS-CoV-2 [19], intervening these infections through regular essential medical services is challenging. Considering the natural history of SARS-CoV-2 infection [20], and high transmissibility from asymptomatic or pre-symptomatic infections [21, 22], capturing these infected individuals is a critical topic. In fact, some universities in the United States have already started to implement a frequent RT-PCR screening test [23-25]. In addition, frequent use of rapid, inexpensive tests, which show lower sensitivity but become positive when the virus load is high, is assumed to be a game changer [26,27], and modelling studies support this presumption [28,29]. While Japan urgently needs to expand the RT-PCR testing capacity for diagnosing patients [11], screening essential workers [12], and raising preparedness for other health emergencies in the near future, using appropriate rapid testing kits for detecting high transmissible cases will be necessary. Moreover, our results indicate that implementing a population-wide screening during a period of circuit breaker will accelerate a decreasing trend of SARS-CoV-2 infection. While this study assumed that only people tested positive would be isolated, because the tracing capacity in Japan has already been overwhelmed, utilizing a population-wide screening as an opportunity for mass isolation and quarantine will aggressively reduce the transmission. In fact, a recent case study from Slovakia [9] suggested that population-wide screening, even conducted only a few times, can be beneficial for detecting infectious individuals, appropriately isolating people who were tested positive, quarantining their close-contacts, and breaking chains of transmission. If managed appropriately, this approach can contribute to minimizing the period of lockdown, thus mitigating the impact of social-distancing policy on socio-economic activities.

Five operational challenges must be noted. First, incentivizing public participation will be exigent. Discrimination against COVID-19 patients and healthcare workers and insufficient capacity in health communication will be eminent barriers [30]. Second, a limited testing capacity and weak logistics in procuring personal protective equipment will be demanding. Adopting a pooling strategy [31-33] will dramatically dilate Japan’s testing capacity and make it possible to repeatedly provide RT-PCR testing. Importing appropriate testing kits for truly recognizing infectiousness based on the previous discussion [34] will be a policy option, although it can be a debatable political choice in terms of health security. Third, the feasibility of mass screening schemes is completely different by context [35]. Discussing advantages and disadvantages of a home-based approach, testing tools, and intervals of mass testing will be vital. While the false-positive result was not a critical matter in population-wide screening in Slovakia [9], it will be sensible if positive cases detected by rapid tests are followed by confirmatory PCR testing. Fourth, our results suggested the necessity of launching shelter hospitals (i.e., isolation facilities). These will not only work for truly breaking chains of transmission and avoiding additional transmissions (e.g., household transmission), but lessen additional burden on tertiary referral hospitals. Fifth, social support, including financial scheme for positive cases, must be ensured with a political will to contain the pandemic. This is crucial to truly achieve a high compliance of isolation. Expanding the public support towards close contacts of patients will improve their compliance.

There are several limitations in this modelling exercise. First, the heterogeneity of COVID-19 infection is not considered. The heterogeneity of the offspring distribution of secondary infection of COVID-19 is recognized [36,37], and intervention in high-risk environments will be effective for preventing superspreading events [38]. We launched this modelling exercise with concerns that a great number of essential workers, even healthcare workers at hospitals and nursing homes, still cannot access to regular protective screening in Japan [12]. Opportunities for population-wide testing can be beneficial for ensuring their access to testing. Second, the management of prefectural borders is not considered. However, as Okinawa prefecture has no land bridge except the U.S. Military Bases and Facilities on Okinawa Island and Its Vicinity, test-based strategy at its prefectural border can be easily implemented compared to other prefectures. Third, our modelling did not strongly address the importance of contact-tracing. While this assumption is plausible in that the tracing capacity in public health centers has been overwhelmed in Japan [11,39], the population-wide testing followed by contact tracing activities, even limited ones, will contribute to further reducing transmission. Fourth, we assumed that the lockdown would decrease social contacts by 50%. Considering that the state of emergency in Japan in April has been declared under the assumption that social contacts will be decreased between 70% and 80% [11], our assumptions can be strengthened.

Despite these limitations, our modelling exercise presented several implications for the deteriorating situation of COVID-19 in Okinawa, Japan. To end this pandemic, revive socio-economic activities and protect citizens’ lives and livelihoods, rigorous investment in public health is manifestly vital [40,41]. Testing obviously plays a vital role in containing the virus, and aggressive, widespread testing can capture more asymptomatic cases. Actually, countries advanced for near-elimination conducted more testing per case [42]. Population-wide screening has been previously practiced for early containment or elimination [7,8], and some countries in Europe started to employ this approach with their own social distancing policies. Our exercise implies that repeated mass testing that targets sufficient population will be a policy option to avoid the lockdown. However, if there are limitations for promptly conducting a mass testing, early introduction of a tight circuit breaker, followed by solid logistics for mass screening, will be critical to contain the pandemic. While expanding the RT-PCR testing capacity and ensuring citizens’ access to testing must be utmost important, critical roles of mass screening must be reconsidered for promptly detecting highly infectious individuals, protecting positive cases, breaking further transmission dynamics, and containing the pandemic.

## Data Availability

Our data are available upon request.

## Acknowledgements

We would like to thank all healthcare workers on the front-line and share their voices with us. We also thank Dr. Makoto Aoki, Dr. Kiyosu Taniguchi, Dr. Taro Kondo and Prof. Kenji Shibuya for sharing their scientific insights.

## References

1. Flaxman S, Mishra S, Gandy A, et al. Estimating the effects of non-pharmaceutical interventions on COVID-19 in Europe. Nature 2020 Aug;584(7820):257–61.

2. Caselli F, Grigoli F, Lian W, Sandri D. The great lockdown: dissecting the economic effects. In: International Monetary Fund. World Economic Outlook, October 2020: A Long and Difficult Ascent. Washington DC: International Monetary Fund, 2020; 65–84.

3. Alvelda P, Ferguson T, Mallery JC. To save the economy, save people first. Available online: https://www.ineteconomics.org/perspectives/blog/to-save-the-economy-save-people-first (Accessed on 10 December 2020).

4. Tokuda Y, Shibuya K, Oguro K. Priority of SARS-CoV-2 test, trace, and isolation in Japan. J Gen Fam Med. 2020 Dec 4: in press.

5. Peto J, Carpenter J, Smith GD, et al. Weekly COVID-19 testing with household quarantine and contact tracing is feasible and would probably end the epidemic. R Soc Open Sci. 2020 Jun 24;7(6):200915.

6. European Centre for Disease Prevention and Control. Population-wide testing of SARS-CoV-2: country experiences and potential approaches in the EU/EEA and the United Kingdom. Available online: https://www.ecdc.europa.eu/sites/default/files/documents/covid-19-population-wide-testing-country-experiences.pdf (accessed on 10 December 2020).

7. Wu Z, Wang Q, Zhao J, et al. Time course of a second outbreak of COVID-19 in Beijing, China, June-July 2020. JAMA 2020 Aug 24;e2015894.

8. Xing Y, Wong GW, Ni W, Hu X, Xing Q. Rapid Response to an Outbreak in Qingdao, China. N Engl J Med. 2020 Dec 3;383(23):e129.

9. Pavelka M, van Zandvoort K, Abbott S, et al. The effectiveness of population-wide, rapid antigen test based screening in reducing SARS-CoV-2 infection prevalence in Slovakia. medRxiv 2020.12.02.20240648.

10. Liverpool City Council. Symptom-free testing. Available online: https://liverpool.gov.uk/masstesting (accessed on 10 December 2020).

11. Shimizu K, Wharton G, Sakamoto H, Mossialos E. Resurgence of covid-19 in Japan. BMJ 2020 Aug 18;370:m3221.

12. Shimizu K, Kondo T, Tokuda Y, Shibuya K. An open letter to Japan’s new Prime Minister. Lancet 2020 Sep 28;396(10259):e57.

13. Kuniya T, Inaba H. Possible effects of mixed prevention strategy for COVID-19 epidemic: massive testing, quarantine and social distancing. AIMS Public Health 2020 Jul 6;7(3):490–503.

14. Linton NM, Kobayashi T, Yang Y, et al. Incubation period and other epidemiological characteristics of 2019 novel coronavirus infections with right truncation: a statistical analysis of publicly available case data. J Clin Med. 2020 Feb 17;9(2):538.

15. Anderson RM, Heesterbeek H, Klinkenberg D, Hollingsworth TD. How will country-based mitigation measures influence the course of the COVID-19 epidemic? Lancet 2020 Mar 21;395(10228):931–4.

16. He X, Lau EHY, Wu P, et al. Temporal dynamics in viral shedding and transmissibility of COVID-19. Nat Med. 2020 May;26(5):672–5.

17. Kuniya T. Prediction of the epidemic peak of coronavirus disease in Japan, 2020. J Clin Med. 2020 Mar 13;9(3):789.

18. The Japan Association for Infectious Diseases. COVID-19 testing and results (in Japanese). Available online: http://www.kansensho.or.jp/uploads/files/topics/2019ncov/covid19_kensakekka_201012.pdf (accessed on 10 December 2020).

19. Buitrago-Garcia D, Egli-Gany D, Counotte MJ, et al. and transmission potential of asymptomatic and presymptomatic SARS-CoV-2 infections: A living systematic review and meta-analysis. PLoS Med. 2020 Sep 22;17(9):e1003346.

20. Sethuraman N, Jeremiah SS, Ryo A. Interpreting diagnostic tests for SARS-CoV-2. JAMA 2020 Jun 9;323(22):2249–51.

21. He X, Lau EHY, Wu P, et al. Author Correction: Temporal dynamics in viral shedding and transmissibility of COVID-19. Nat Med. 2020 Sep;26(9):1491–3.

22. Furukawa NW, Brooks JT, Sobel J. Evidence supporting transmission of severe acute respiratory syndrome coronavirus 2 while presymptomatic or asymptomatic. Emerg Infect Dis. 2020 Jul;26(7):e201595.

23. Boston University. Back to Boston University: COVID-19 Screening, Testing & Contact Tracing. Available online: https://www.bu.edu/back2bu/student-health-safety/covid-19-screening-testing-contact-tracing/ (accessed on 10 December 2020).

24. Harvard University. Testing & Tracing. Available online: https://www.harvard.edu/coronavirus/testing-tracing (accessed on 10 December 2020).

25. MIT Covid Apps. COVID-19 testing requirements and procedures. Available online: https://covidapps.mit.edu/medical-testing-information (accessed on 10 December 2020).

26. Mina MJ, Parker R, Larremore DB. Rethinking covid-19 test sensitivity—A strategy for containment. N Engl J Med. 2020 Nov 26;383(22):e120.

27. Yamey G, Walensly RP. Covid-19: re-opening universities is high risk. BMJ 2020 Sep 1;370:m3365.

28. Paltiel AD, Zheng A, Walensky RP. Assessment of SARS-CoV-2 Screening Strategies to Permit the Safe Reopening of College Campuses in the United States. JAMA Netw Open 2020 Jul 1;3(7):e2016818.

29. Larremore DB, Wilder B, Lester E, et al. Test sensitivity is secondary to frequency and turnaround time for COVID-19 screening. Sci Adv. 2020 Nov 20;eabd5393.

30. Shimizu K, Lin L. Defamation against healthcare workers during COVID-19 pandemic. Int J Health Policy Manag. 2020 Sep 27: in press.

31. Hirotsu Y, Maejima M, Shibusawa M, et al. Pooling RT-qPCR testing for SARS-CoV-2 in 1000 individuals of healthy and infection-suspected patients. Sci Rep. 2020 Nov 3;10(1):18899.

32. Mutesa L, Ndishimye P, Butera Y, et al. A pooled testing strategy for identifying SARS-CoV-2 at low prevalence. Nature 2020 Oct 21: in press.

33. Taylor L. Uruguay is winning against covid-19. This is how. BMJ 2020 Sep 18;370:m3575.

34. Rubin R. The Challenges of Expanding Rapid Tests to Curb COVID-19. JAMA 2020 Nov 10;324(18):1813–5.

35. Mckee M, Nagyova I. Could Slovakia’s mass testing programme work in England? Available online: https://blogs.bmj.com/bmj/2020/12/07/could-slovakias-mass-testing-programme-work-in-england/ (accessed on 10 December 2020).

36. Shi Q, Hu Y, Peng B, et al. Effective control of SARS-CoV-2 transmission in Wanzhou, China. Nat Med. 2020 Nov 30: in press.

37. Sun K, Wang W, Gao L, et al. Transmission heterogeneities, kinetics, and controllability of SARS-CoV-2. Science 2020 Nov 24:eabe2424.

38. Chang S, Pierson E, Koh PW, et al. Mobility network models of COVID-19 explain inequities and inform reopening. Nature 2020 Nov 10: in press.

39. Shimizu K, Negita M. Lessons Learned from Japan’s Response to the First Wave of COVID-19: A Content Analysis. Healthcare (Basel) 2020 Oct 23;8(4):426.

40. Walensky RP, Rio CD. From Mitigation to Containment of the COVID-19 Pandemic: Putting the SARS-CoV-2 Genie Back in the Bottle. JAMA 2020 May 19;323(19):1889–90.

41. Cutler DM, Summers LH. The COVID-19 Pandemic and the $16 Trillion Virus. JAMA 2020 Oct 20;324(15):1495–96.

42. Rannan-Eliya RP, Wijemunige N, Gunawardana J, et al. Increased Intensity Of PCR Testing Reduced COVID-19 Transmission Within Countries During The First Pandemic Wave. Health Aff. (Millwood) 2020 Dec 2;101377hlthaff202001409.

